# Causal effect of adiposity on the risk of 19 gastrointestinal diseases: a Mendelian randomization study

**DOI:** 10.1101/2021.11.19.21266578

**Authors:** Min Seo Kim, Minku Song, Soyeon Kim, Beomsu Kim, Wonseok Kang, Jong Yeob Kim, Woojae Myung, Inhyeok Lee, Ron Do, Amit V Khera, Hong-Hee Won

**Author notes:** Correspondence to: Hong-Hee Won, PhD, Samsung Advanced Institute for Health Sciences & Technology (SAIHST), Sungkyunkwan University, Samsung Medical Center, Seoul, Republic of Korea. These authors contributed equally to this work (Min Seo Kim, Minku Song). This manuscript has been reviewed and approved by all authors. Contributors: MS Kim, M Song, and H Won contributed to the concept and design of this study. MS Kim, M Song, and H Won analyzed and interpreted the data. S Kim, B Kim, and R Do provided statistical and methodological supervision of the study. MS Kim and M Song wrote the first draft of the manuscript. S Kim, B Kim, W Kang, JY Kim, W Myung, I Lee, R Do, and AV Khera contributed to the data interpretation and critical revision of the manuscript. All authors have read and approved the final version of the manuscript. Disclosure: All authors have completed the ICMJE uniform disclosure form at www.icmje.org/coi_disclosure.pdf (available on request from the corresponding author) and declare the following: no support from any organization for the submitted work; no financial relationships with any organizations that might have an interest in the submitted work in the previous three years; no other relationships or activities that could appear to have influenced the submitted work.

## Abstract

**Objectives:** We applied Mendelian randomization (MR) to investigate the causal associations of body mass index (BMI) and waist circumference (WC) with 19 gastrointestinal (GI) disorders.

**Design:** MR study.

**Setting:** The UK Biobank, Genetic Investigation of Anthropometric Traits (GIANT) Consortium, FinnGen consortium, and genome-wide association studies.

**Participants:** Overall, >400,000 UK Biobank participants, >170,000 participants of Finnish descent, and numerous consortia participants with predominantly European ancestry.

**Interventions:** Single-nucleotide polymorphisms associated with BMI and WC were used as instrumental variables to estimate the causal associations with the GI conditions.

**Main outcome measures:** Risk of developing 19 GI diseases

**Results:** After correction for multiple testing (Bonferroni-corrected threshold of P<0.05/19) and testing for consistencies using several MR methods with varying assumptions (inverse variance weighted, weighted median, MR-Egger, and MR-PRESSO), genetically predicted BMI was associated with increased risks of non-alcoholic fatty liver disease (NAFLD), cholecystitis, cholelithiasis, and primary biliary cholangitis. The odds ratio (OR) per one standard deviation (SD) increased in genetically predicted BMI (4.77 kg/m^2^) from 1.22 (95% confidence interval [CI] 1.12–1.34; P<0.0001) for NAFLD to 1.65 (95% CI 1.31–2.06; P<0.0001) for cholecystitis. Genetically predicted WC was associated with increased risks of NAFLD, alcoholic liver disease (ALD), cholecystitis, cholelithiasis, colon cancer, and gastric cancer. ALD was associated with WC even after adjustment for alcohol consumption in multivariable MR analysis. The OR per 1 SD increased in genetically predicted WC (12.52 cm) from 1.41 (95% CI 1.17–1.70; P=0.0015) for gastric cancer to 1.74 (95% CI 1.21–1.78; P<0.0001) for cholelithiasis.

**Conclusions:** Higher BMI and WC are causally associated with an increased risk of GI abnormalities, particularly of hepatobiliary organs (liver, biliary tract, and gallbladder) that are functionally related to fat metabolism. Abdominal obesity measured by WC might be more influential and relevant with a diverse span of GI diseases than BMI, highlighting a possible pathophysiological role of visceral abdominal fats in the development of GI disorders and cancers.

**Graphical Abstract:** 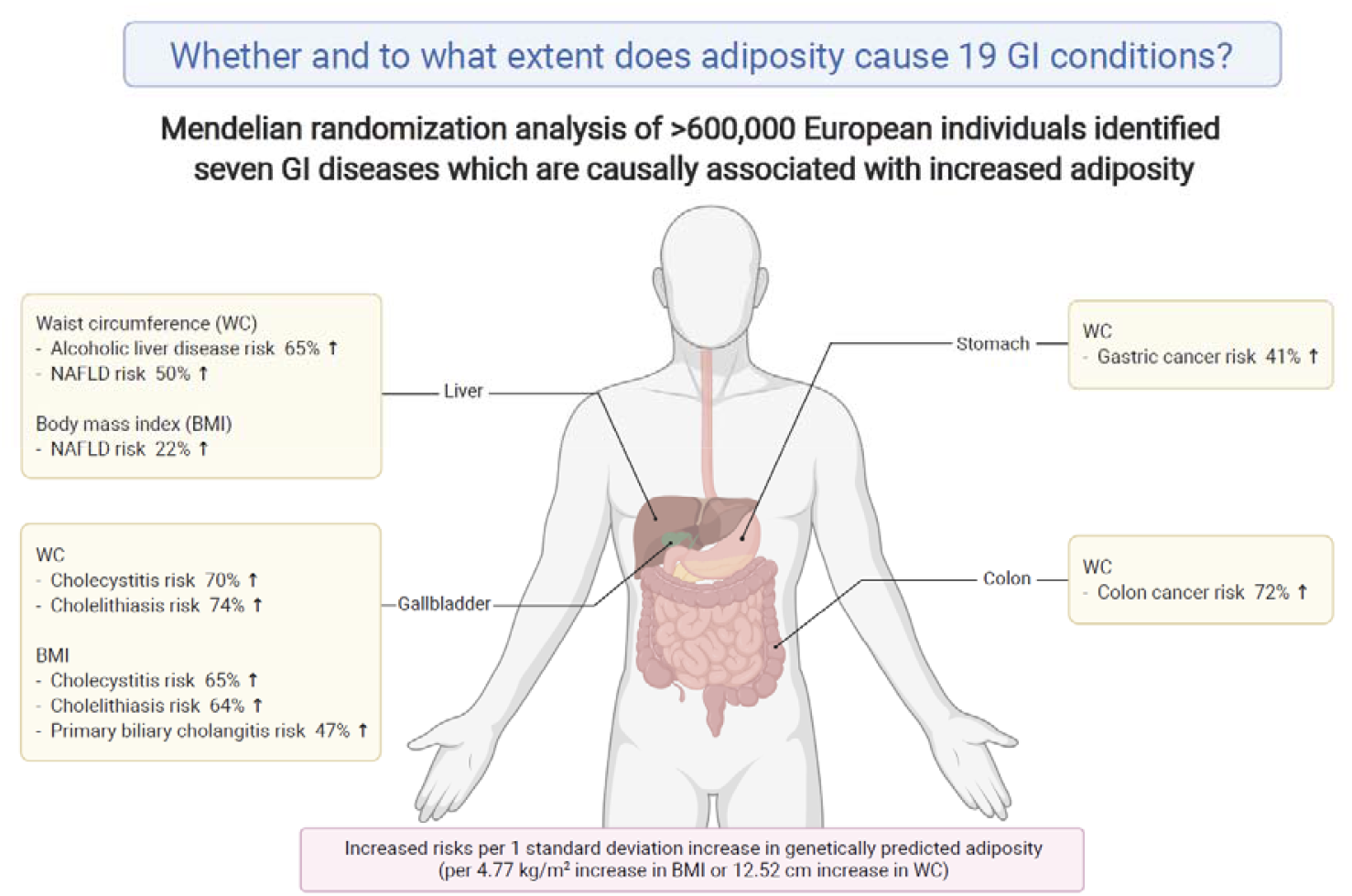

## Introduction

Global anthropometric metrics have rapidly changed from predominantly normal weight or underweight to overweight or obese in the past few decades.^1^ These immense changes in global adiposity status over a relatively short period are likely to be driven by changes in global diet to a more processed, affordable, and high-sugar food, alongside the sedentary lifestyle associated with automation of many workloads and commuting modes.^2^ With the steep increase in the number of people who are obese worldwide, especially in low- and middle-income countries,^3^ it has become increasingly important to identify the potential impact of fat mass quantity and its distribution on human health. Thus far, the relationship between adiposity and cardiovascular diseases has been actively investigated,^4,5^ whereas the relationship between adiposity and gastrointestinal (GI) outcomes has been relatively unvisited. Despite the abdomen being a predominant place where fats are concentrated, the adjacent GI organs that are likely subject to the paracrine effect of visceral fat-originated cytokines were paradoxically overlooked.

Although numerous cross-sectional and cohort studies have explored the relationship between adiposity and GI outcomes,^6-8^ they are observational and provide evidence of association but not causation.^9^ Causation could be inferred from randomized controlled trial (RCT) designs, but interventions inducing obesity are neither practical nor ethical, limiting the elucidation of causal inference. With limited evidence from observational and interventional studies, the human genetics approach, Mendelian randomization (MR), offers an opportunity to reliably inspect the potential causal effect between increased adiposity and multiple GI phenotypes.^10^ MR approach is based on the concept of the naturally occurring random allocation of alleles during meiosis and uses genetic variants associated with risk factors as instrumental variables. This process is conceptually analogous to the random allocation of participants in an RCT.^10,11^ MR can overcome biases such as reverse causation and confounding, of which observational studies are particularly susceptible.^12^ Employing this well established genetic study design,^10,13^ we conducted an MR analysis to examine the causal relationship between adiposity and the risk of 19 GI disorders.

## Methods

### Study design

This study was designed to scrutinize the causal effect of adiposity on the risk of multiple GI outcomes (Figure 1). We used an MR approach in which genetic variants associated with a risk factor (or exposure) were used as instrumental variables to infer causality of the risk factor on the outcome of interest.^12^ MR analyses limit biases induced by confounding and reverse causation, which has long hampered observational studies to elucidate precise measures for epidemiological relationships.^10^ We used two-sample Mendelian randomization (TSMR) and utilized summary statistics from relevant genome-wide association studies (GWAS) of exposures and outcomes. Compared to the one-sample MR approach, TSMR is known to be less vulnerable to the winner’s curse and weak instrument bias.^14^ Moreover, TSMR has increased statistical power by using summary data obtained from large-scale GWAS consortia.^15^ We purposefully avoided using dependent and overlapping samples for exposure and outcome data sets to prevent inflation of genetic association statistics. We further matched for ethnicity to minimize population stratification bias.^14,16^ For GI outcomes, we used summary statistics from studies with appropriate case-control balance or those controlling case-control imbalance using methods such as Scalable and Accurate Implementation of GEneralized mixed model (SAIGE) to minimize inflated type 1 error due to extreme case-control imbalance.^17^

**Figure 1.**
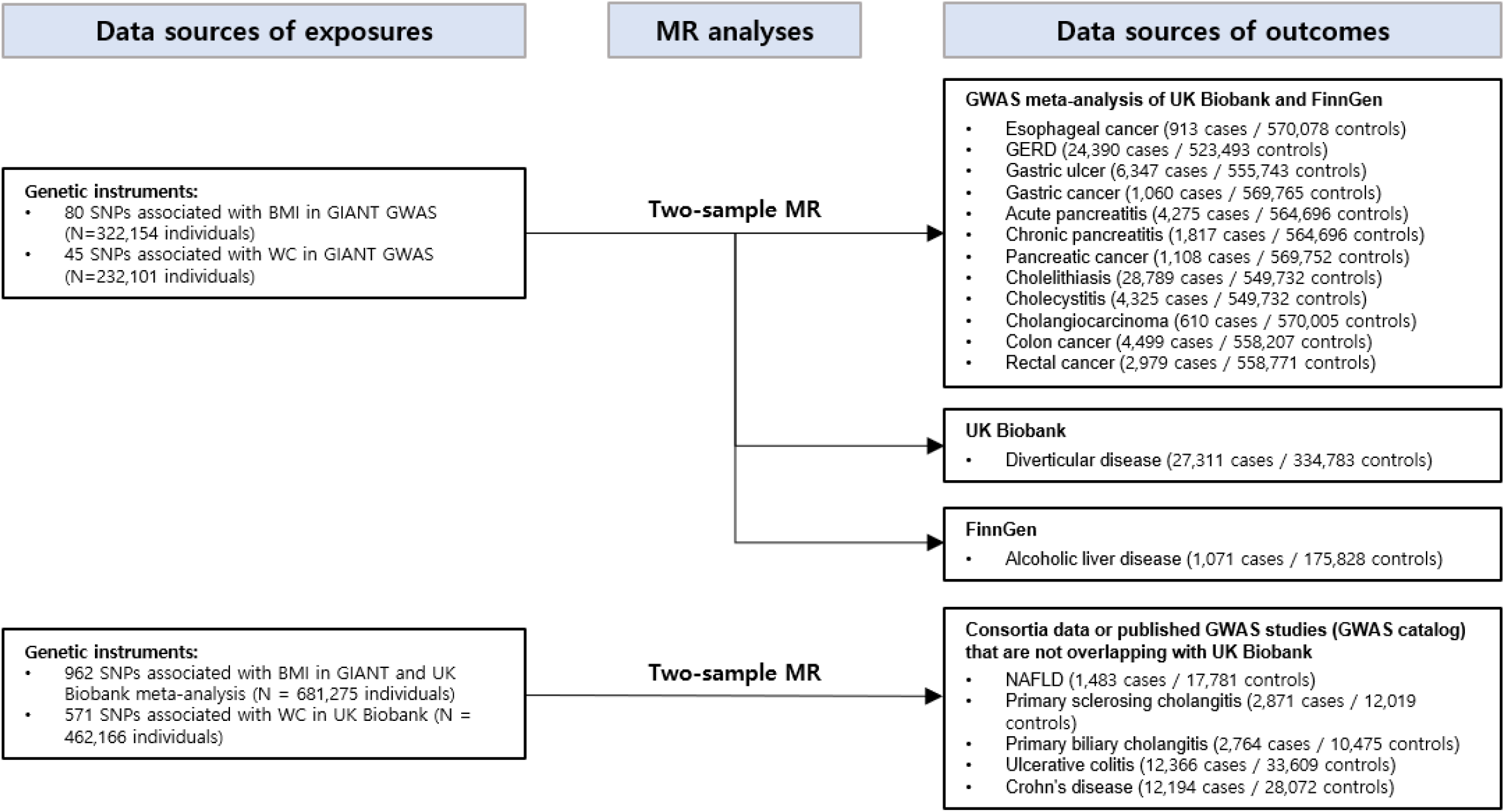
Study flow diagram and data sources used in this study. SNP, single-nucleotide polymorphism; GIANT, Genetic Investigation of Anthropometric Traits consortium; GWAS, genome-wide association study; GERD, gastroesophageal reflux disease; MR, Mendelian randomization; NAFLD, non-alcoholic fatty liver disease.

### Data for exposure

We leveraged data of single-nucleotide polymorphisms (SNPs) that are associated with the adiposity at genome-wide significant levels (P<5 × 10^−8^) discovered by GWAS from two large cohorts: Genetic Investigation of Anthropometric Traits (GIANT) Consortium^18-21^ and UK Biobank.^17,22^ Two major adiposity measuring indices, body mass index (BMI) and waist circumference (WC), were used to account for the varying effects of fat distribution on clinical GI outcomes (Supplementary Table 2). BMI has traditionally been used to measure body size,^23^ while WC was assessed, more specifically, to indicate abdominal or central obesity.^24^ We investigated these two measures simultaneously following the consensus statement from the International Atherosclerosis Society (IAS) and International Chair on Cardiometabolic Risk (ICCR) Working Group on Visceral Obesity, arguing the necessity of using both BMI and WC.^24^ We did not utilize other measures that indicate fat distributions such as visceral adipose tissue volume, arm-fat-ratio, leg-fat-ratio, and trunk-fat-ratio considering that the summary statistics were derived from mixed ancestry populations or studies including UK Biobank participants.^25,26^ The summary data from the GIANT consortium and UK Biobank used in this study were adjusted for age, sex, and major genetic principal components. We selected independent SNPs using linkage disequilibrium clumping with an *r*^2^ threshold of 0.01 and clumping window within 10,000 kb using the 1000 Genomes Project reference panel (European population), as performed in previous studies.^27-30^

### Data for outcome

We investigated 19 GI diseases that have been analyzed in GWAS and thus have relevant information of genetic variants associated with the diseases. Diseases of GWAS with a low number of cases (<500) were excluded from our analysis (Barrett’s esophagus, diverticulum of esophagus, hepatocellular carcinoma, other liver cancers, and small intestinal cancers) as they are at high risk of false inference due to limited statistical power. Chronic liver diseases (CLD) and cirrhosis were excluded as most hepatobiliary conditions could eventually progress to CLD in the end stage. Therefore, disease-specific inferences could not be drawn for CLD and cirrhosis. The diseases included were gastroesophageal reflux disease (GERD), esophageal cancer, gastric ulcer, gastric cancer, colon cancer, rectal cancer, acute pancreatitis, chronic pancreatitis, pancreatic cancer, cholelithiasis, cholecystitis, cholangiocarcinoma, chronic liver disease/cirrhosis, non-alcoholic fatty liver disease (NAFLD), alcoholic liver disease (ALD), primary sclerosing cholangitis (PSC), primary biliary cholangitis (PBC), diverticular diseases, ulcerative colitis (UC), and Crohn’s disease (CD) (Supplementary Table 1). The genetic variants associated with the 19 GI outcomes were identified from: (1) UK Biobank data, a large prospective cohort study with more than 500,000 people aged 40-69 years during recruitment between 2006 and 2010, (2) R4 release of the FinnGen consortium data, including up to 170,000 participants of Finnish descent, and (3) GWAS catalog data provided by the National Human Genome Research Institute (NHGRI) and the European Bioinformatics Institute (EMBL-EBI). For available outcomes, we meta-analyzed non-overlapping cohorts to increase the statistical power.^31,32^ When applicable, UK Biobank and FinnGen summary statistics were combined using METAL, a fixed-effects inverse variance method, based on the beta effect estimates, standard errors, and P-values of the SNPs. The UK Biobank study was approved by the North West Multi-center Research Ethics Committee (MREC) and National Information Governance Board for Health and Social Care (NIGB). FinnGen’s recruitment protocols followed the biobank protocols approved by Fimea, the National Supervisory Authority for Welfare and Health. The FinnGen study was approved by The Coordinating Ethics Committee of the Hospital District of Helsinki and Uusimaa.^33^

### Statistical analysis

We used four MR methods (inverse variance weighted [random-effects model], weighted median, MR-Egger, and MR-PRESSO) to test the consistency of the results across varying assumptions of heterogeneity and pleiotropy effects. The inverse variance weighted (IVW) method assumes that all genetic variants are valid, but this method is prone to bias when a large portion of SNPs is subject to horizontal pleiotropy.^34^ The weighted median method assumes that most genetic variants are valid. This method is suitable if the proportion of horizontal pleiotropic variants is <50%.^35^ When most genetic variants have horizontal pleiotropy (>50%), the MR-Egger method performs well to test causal estimations, despite being lower in statistical power than other methods.^36^ The MR-PRESSO method was used to test whether horizontal pleiotropic outliers cause bias in the IVW test and exclude outlier SNPs from analysis to draw a more balanced and reliable causal estimation.^37^ To minimize the risk of false-positive results and provide proper interpretation, we deemed the causal association ‘robust’ when at least three of the aforementioned methods presented consistent results in terms of effect direction and statistical significance and the P-value reached the Bonferroni-corrected threshold (P<0.0026, alpha=0.05/19), as demonstrated in a previous study.^38^ However, given that inconsistency between the robust methods does not necessarily exclude the possibility of the relationship being genuinely causal,^14^ we have not precluded inconsistent results from suggestive causal inferences.

We performed multivariable MR analyses adjusting for well established risk factors of GI problems, including alcohol consumption and smoking.^39-41^ We used the IVW method to overcome potential confounding and estimate the causal effect of multiple exposure variables independently on outcomes.^42-44^ The multivariable MR is useful in estimating a direct causal effect when two or more variables are correlated with the exposure of interest, such as when both exposures may exert a causal effect on the outcome or when one may mediate the effect of the other on the outcome. For certain relationships where the risk for reverse causation would be high due to a lack of previous evidence ensuring the direction of effect (i.e., PBC/PSC and UC/CD), we performed a bidirectional MR analysis to clarify the direction of effect. We also performed a leave-one-out analysis to identify if a single genetic variant strongly drove the effect of the association.

In forest plots, we presented the best causal estimations for each GI outcome considering both pleiotropy and heterogeneity effect.^38^ A detailed flow diagram for selecting the best causal estimation is described in Supplementary Figure 1. We assessed heterogeneity using Cochran’s Q-test and I^2^ and visualized them using scatter plots.^45^ Unbalanced pleiotropy was inspected using Egger regression intercept, and horizontal pleiotropy was investigated using a global test of MR-PRESSO.^45^ The statistical power of MR was calculated using the previously established method proposed by Brion *et al*.^46^ The statistical power > 80% was deemed acceptable to assess the effect of adiposity on GI.^47-49^ We applied Bonferroni’s correction (P<0.0026, based on alpha=0.05/19 GI outcomes) to stringently correct for multiple testing.^50^ Associations with a P-value between 0.0026 and 0.05 were deemed suggestively causal inference regardless of sensitivity analyses. Associations were considered robust only when they reached the Bonferroni-corrected threshold (P<0.0026, alpha=0.05/19), and three out of four MR methods (inverse variance weighted, weighted median, MR-Egger, and MR-PRESSO) showed consistent results. All statistical analyses were carried out using R version 4.0.4 (R Foundation) and R package (‘TwoSampleMR’ and ‘MendelianRandomization’) for the MR analysis.^51,52^

## Results

### Main findings

The data sources of the outcomes (i.e., GIANT, UK Biobank, and multiple consortia) are provided in Supplementary Table 1. The 80 BMI-associated SNPs explained 1.61% of the variance in BMI (corresponding to an F statistic of 66) when only GIANT GWAS data were used. In contrast, the 962 BMI-associated SNPs explained 7.85% of the variance in BMI (F statistic, 60.3) when GIANT and UK Biobank GWAS data were meta-analyzed (for the analysis of GI outcomes from consortia composed of those not overlapping with GIANT and UK Biobank participants). The 45 WC-associated SNPs from GIANT GWAS data explained 1.14% of the variance in WC (F statistic of 59.9) (Supplementary Table 2). Statistical powers for GI outcomes are provided in Supplementary Tables 3 and 4.

After correction for multiple testing (Bonferroni-corrected threshold of P<0.0026) and testing for consistencies across the four MR methods (IVW, weighted median, MR-Egger, and MR-PRESSO), genetically predicted BMI was robustly associated with increased risks of NAFLD, cholecystitis, cholelithiasis, and PBC. For the robust associations, the odds ratio (OR) per 1 standard deviation (SD) increase in BMI ranged from 1.22 (95% confidence interval (CI) 1.12–1.34; P<0.0001) for NAFLD to 1.65 (95% CI 1.31–2.06; P<0.0001) for cholecystitis (Figure 2). After correction for multiple testing and sensitivity analysis, genetically predicted WC was robustly associated with increased risks of NAFLD, ALD, cholecystitis, cholelithiasis, colon cancer, and gastric cancer. The OR per 1 SD increase in WC for such robust associations ranged from 1.41 (95% CI 1.17–1.70; P=0.0015) for gastric cancer to 1.74 (95% CI 1.21–1.78; P<0.0001) for cholelithiasis (Figure 3).

**Figure 2.**
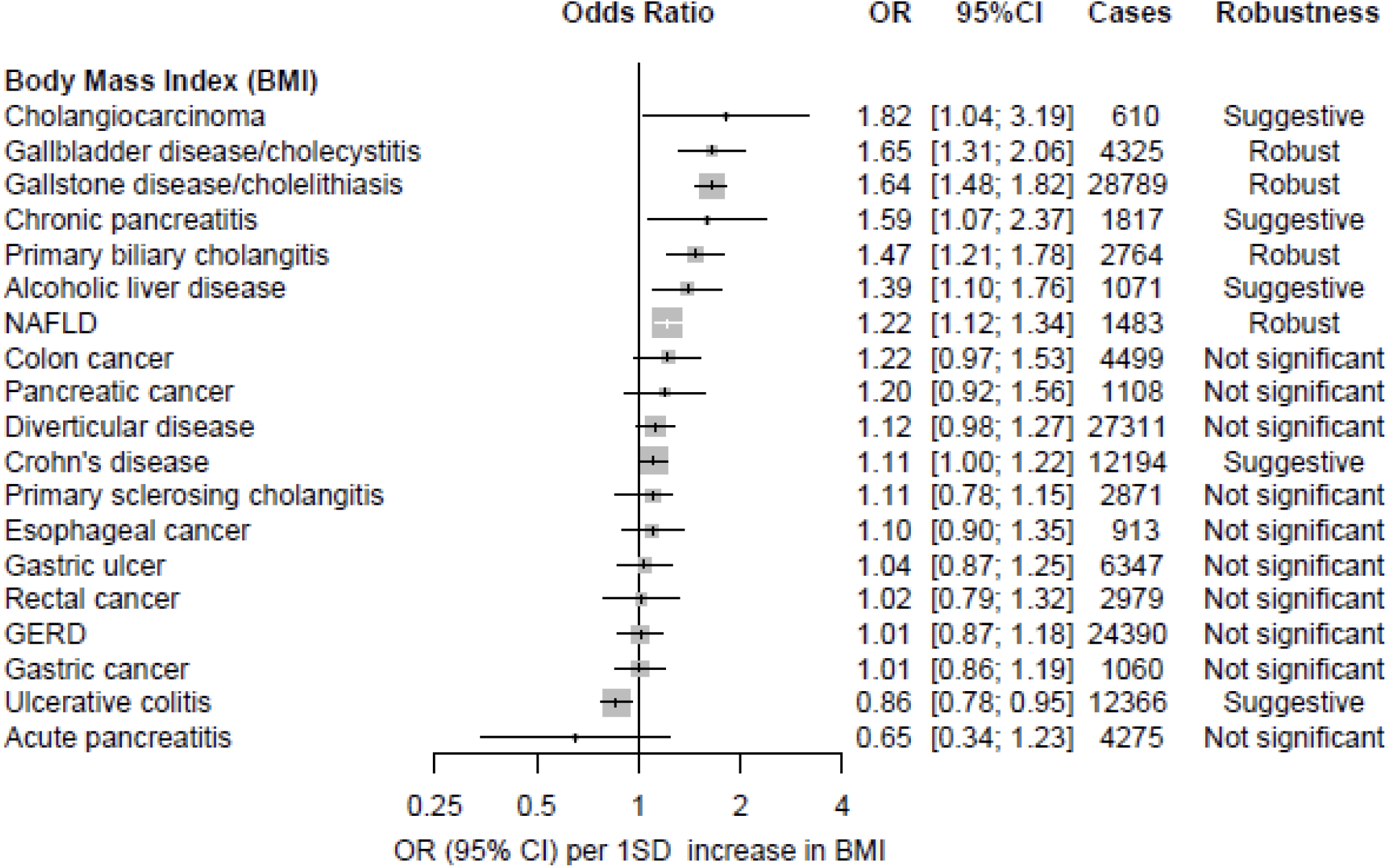
Associations of genetically predicted one standard deviation (SD) increase in body mass index (4.77 kg/m^2^) with 19 gastrointestinal conditions. The robustness of the results was judged when the Bonferroni-corrected threshold was reached (P<0.0026, alpha = 0.05/19), and three out of four MR methods (inverse variance weighted, weighted median, MR-Egger, and MR-PRESSO) showed consistent results. The forest plot presents the best causal estimate for each GI outcome. While numerous phenotypes were suggestively causative (P<0.05), only four outcomes (cholecystitis, cholelithiasis, PBC, and NAFLD) remained robust after multiple testing and sensitivity analyses across diverse MR methods. CI, confidence interval; GERD, gastroesophageal reflux disease; NAFLD, non-alcoholic fatty liver disease.

**Figure 3.**
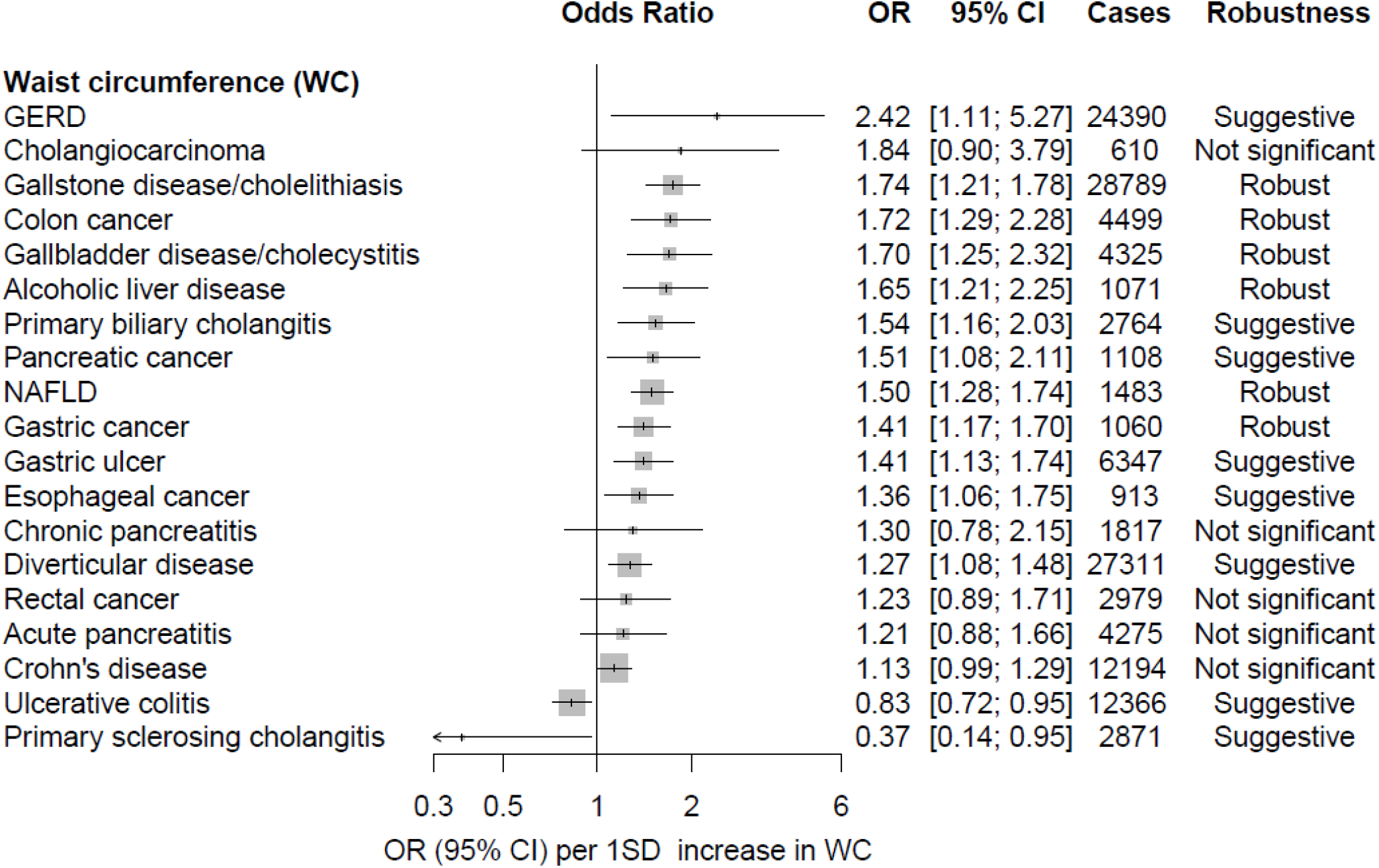
Associations of genetically predicted one standard deviation (SD) increase in waist circumference (12.52 cm) with 19 gastrointestinal conditions. The robustness of the results was judged when the Bonferroni-corrected threshold was reached (P<0.0026, alpha = 0.05/19), and three out of four MR methods (inverse variance weighted, weighted median, MR-Egger, and MR-PRESSO) showed consistent results. The forest plot presents the best causal estimate for each GI outcome. While numerous phenotypes were suggestively causative (P<0.05), six outcomes (cholecystitis, cholelithiasis, NAFLD, alcoholic liver disease, colon cancer, and gastric cancer) remained robust after multiple testing and sensitivity analyses across diverse MR methods. CI, confidence interval; GERD, gastroesophageal reflux disease; NAFLD, non-alcoholic fatty liver disease.

MR analyses showed that genetically predicted BMI was suggestively associated (P<0.05) with increased risks of cholangiocarcinoma, cholecystitis, cholelithiasis, chronic pancreatitis, PBC, ALD, NAFLD, diverticular disease, CD, and the decreased risk of UC (Table 1). Genetically predicted WC was suggestively associated (P<0.05) with the increased risks of GERD, cholelithiasis, cholecystitis, colon cancer, ALD, PBC, pancreatic cancer, NAFLD, gastric cancer, esophageal cancer, diverticular disease, and decreased risks of UC and PSC (Table 2).

**Table 1.** Two-sample Mendelian randomization causal estimations for the effect of body mass index (BMI) on 19 gastrointestinal outcomes

| Outcome | Inverse variance weighted | | Weighted median | | MR-Egger | | MR-Egger intercept | | $P_h$ | Q statistics | MR-PRESSO (outlier corrected) | | P horizontal pleiotropy | P distortion | Outlier SNPs |
| --- | --- | --- | --- | --- | --- | --- | --- | --- | --- | --- | --- | --- | --- | --- | --- |
|  | OR (95% CI) | P value | OR (95% CI) | P value | OR (95% CI) | P value | Egger intercept | P value |  |  | OR (95% CI) | P value |  |  |  |
| GERD | **1.01**<br>(0.87-1.18) | **8.79E-01** | 1.06<br>(0.84-1.33) | 6.40E-01 | 1.57<br>(0.99-2.48) | 5.82E-02 | -0.01250 | 5.04E-02 | 5.27E-01 | 72 | - | - | - | - | - |
| Colon cancer | 1.17<br>(0.92-1.49) | 1.92E-01 | 1.30<br>(0.95-1.8) | 1.03E-01 | 1.29<br>(0.64-2.61) | 4.79E-01 | -0.00281 | 7.74E-01 | 3.90E-02 | 73 | **1.22**<br>(0.97-1.53) | **8.79E-02** | 0.042 | 0.772 | 1 |
| Acute pancreatitis | 1.24<br>(0.99-1.55) | 5.70E-02 | 1.22<br>(0.87-1.71) | 2.44E-01 | **0.65**<br>(0.34-1.23) | **1.92E-01** | 0.01850 | 3.91E-02 | 3.69E-01 | 73 | - | - | - | - | - |
| Chronic pancreatitis | **1.59**<br>(1.07-2.37) | **2.19E-02** | 1.25<br>(0.71-2.2) | 4.42E-01 | 1.59<br>(0.49-5.2) | 4.45E-01 | 0.00005 | 9.98E-01 | 8.89E-01 | 72 | - | - | - | - | - |
| Cholelithiasis | **1.64**<br>(1.48-1.82) | **1.44E-20** | 1.64<br>(1.42-1.89) | 8.78E-12 | 1.85<br>(1.36-2.52) | 1.97E-04 | -0.00351 | 4.14E-01 | 7.75E-02 | 73 | - | - | - | - | - |
| Pancreatic cancer | 1.08<br>(0.86-1.36) | 5.03E-01 | **1.20**<br>(0.92-1.56) | **1.85E-01** | 1.71<br>(0.88-3.33) | 1.20E-01 | -0.01320 | 1.57E-01 | 4.45E-06 | 73 | - | - | - | - | - |
| Gastric ulcer | 1.10<br>(0.89-1.35) | 3.84E-01 | 1.25<br>(1.02-1.53) | 3.17E-02 | 1.57<br>(0.85-2.9) | 1.51E-01 | -0.01040 | 2.24E-01 | 7.72E-28 | 73 | **1.04**<br>(0.87-1.25) | **6.57E-01** | <0.001 | 0.349 | 6 |
| Rectal cancer | **1.02**<br>(0.79-1.32) | **8.77E-01** | 0.90<br>(0.61-1.32) | 5.74E-01 | 1.86<br>(0.88-3.94) | 1.10E-01 | -0.01730 | 1.00E-01 | 6.34E-01 | 73 | - | - | - | - | - |
| Esophageal cancer | 1.10<br>(0.88-1.38) | 4.11E-01 | 1.23<br>(0.97-1.56) | 8.81E-02 | 1.72<br>(0.89-3.34) | 1.11E-01 | -0.01300 | 1.61E-01 | 1.04E-16 | 73 | **1.10**<br>(0.9-1.35) | **3.58E-01** | <0.001 | 0.979 | 4 |
| Gastric cancer | 1.09<br>(0.9-1.33) | 3.73E-01 | 1.20<br>(1.01-1.43) | 3.37E-02 | 1.49<br>(0.83-2.68) | 1.81E-01 | -0.00898 | 2.69E-01 | 1.30E-40 | 73 | **1.01**<br>(0.86-1.19) | **9.14E-01** | <0.001 | 0.184 | 11 |
| Cholecystitis | **1.65**<br>(1.31-2.06) | **1.48E-05** | 1.41<br>(1.01-1.97) | 4.08E-02 | 1.31<br>(0.67-2.55) | 4.28E-01 | 0.00656 | 4.79E-01 | 2.10E-01 | 73 | - | - | - | - | - |
| Cholangiocarcinoma | **1.82**<br>(1.04-3.19) | **3.60E-02** | 1.29<br>(0.56-2.99) | 5.55E-01 | 0.62<br>(0.12-3.24) | 5.74E-01 | 0.03100 | 1.79E-01 | 7.52E-01 | 73 | - | - | - | - | - |
| Diverticular disease | 1.11<br>(0.97-1.28) | 1.22E-01 | 1.12<br>(0.97-1.3) | 1.34E-01 | 0.80<br>(0.54-1.19) | 2.71E-01 | 0.00964 | 8.47E-02 | 1.21E-09 | 73 | **1.12**<br>(0.98-1.27) | **9.90E-02** | <0.001 | 0.824 | 3 |
| Ulcerative colitis | 0.85<br>(0.76-0.95) | 3.13E-03 | 0.89<br>(0.77-1.03) | 1.22E-01 | 0.98<br>(0.73-1.33) | 9.18E-01 | -0.00235 | 3.13E-01 | 2.92E-33 | 814 | **0.86**<br>(0.78-0.95) | **2.53E-03** | <0.001 | 0.889 | 16 |
| Crohn's disease | 1.14<br>(1.01-1.28) | 3.14E-02 | 1.27<br>(1.09-1.47) | 1.69E-03 | 1.56<br>(1.12-2.16) | 8.01E-03 | -0.00507 | 4.32E-02 | 2.22E-51 | 814 | **1.11**<br>(1-1.22) | **4.17E-02** | <0.001 | 0.597 | 23 |
| NAFLD | 1.22<br>(1.11-1.34) | 4.62E-05 | 1.02<br>(0.91-1.15) | 7.63E-01 | 1.50<br>(1.04-2.18) | 3.09E-02 | -0.00321 | 2.47E-01 | 2.15E-13 | 830 | **1.22**<br>(1.12-1.34) | **1.49E-05** | <0.001 | 0.97 | 4 |
| ALD | **1.39**<br>(1.1-1.76) | **5.64E-03** | 1.56<br>(1.03-2.36) | 3.63E-02 | 1.09<br>(0.56-2.13) | 8.02E-01 | 0.00393 | 4.42E-01 | 1.40E-01 | 822 | - | - | - | - | - |
| Primary sclerosing cholangitis | 0.91<br>(0.75-1.12) | 3.87E-01 | 1.00<br>(0.73-1.36) | 9.92E-01 | 0.68<br>(0.38-1.24) | 2.10E-01 | 0.00461 | 3.08E-01 | 9.55E-04 | 613 | **1.11**<br>(0.78-1.15) | **5.83E-01** | <0.001 | 0.425 | 4 |
| Primary biliary cholangitis | 1.42<br>(1.16-1.73) | 6.26E-04 | 1.51<br>(1.08-2.11) | 1.65E-02 | 1.80<br>(1.02-3.17) | 4.19E-02 | -0.00382 | 3.76E-01 | 2.91E-06 | 720 | **1.47**<br>(1.21-1.78) | **1.08E-04** | <0.001 | 0.682 | 4 |

The best causal estimation highlighted in bold. Detailed flow diagram for selecting the best causal estimation is described in Supplementary figure 1.
Odds ratio per 1 standard deviation increase in genetically predicted BMI (4.77 kg/m<sup>2</sup>).
The blank (-) for MR-PRESSO estimation indicates no significant outliers.
SNP, single-nucleotide polymorphism; GERD, gastroesophageal reflux disease; NAFLD, non-alcoholic fatty liver disease; ALD, alcoholic liver disease; OR, odds ratio; CI, confidence interval; $P_h$ , P value for heterogeneity.

**Table 2.** Two-sample Mendelian randomization causal estimations for the effect of waist circumference (WC) on 19 gastrointestinal outcomes

| Outcome | Inverse variance weighted | | Weighted median | | MR-Egger | | MR-Egger intercept | | $P_h$ | Q statistics | MR-PRESSO (outlier corrected) | | P horizontal pleiotropy | P distortion | Outlier SNPs |
| --- | --- | --- | --- | --- | --- | --- | --- | --- | --- | --- | --- | --- | --- | --- | --- |
|  | OR (95% CI) | P value | OR (95% CI) | P value | OR (95% CI) | P value | Egger intercept | P value |  |  | OR (95% CI) | P value |  |  |  |
| GERD | 1.01 (0.8-1.28) | 9.27E-01 | 0.99 (0.72-1.36) | 9.67E-01 | **2.42 (1.11-5.27)** | **3.22E-02** | -0.02479 | 2.75E-02 | 3.31E-02 | 40 | - | - | - | - | - |
| Colon cancer | **1.72 (1.29-2.28)** | **1.98E-04** | 1.79 (1.19-2.7) | 5.13E-03 | 1.22 (0.45-3.3) | 7.03E-01 | 0.00982 | 4.83E-01 | 2.75E-01 | 40 | - | - | - | - | - |
| Acute pancreatitis | **1.21 (0.88-1.66)** | **2.39E-01** | 1.07 (0.7-1.64) | 7.62E-01 | 0.62 (0.21-1.85) | 3.96E-01 | 0.01897 | 2.18E-01 | 1.01E-01 | 40 | - | - | - | - | - |
| Chronic pancreatitis | **1.30 (0.78-2.15)** | **3.09E-01** | 1.05 (0.52-2.12) | 8.95E-01 | 3.22 (0.55-18.77) | 2.00E-01 | -0.02582 | 2.98E-01 | 8.36E-01 | 40 | - | - | - | - | - |
| Cholelithiasis | 1.81 (1.54-2.11) | 1.67E-13 | 1.74 (1.44-2.1) | 5.81E-09 | 2.79 (1.64-4.77) | 5.42E-04 | -0.01238 | 1.04E-01 | 3.46E-03 | 40 | **1.74 (1.21-1.78)** | **1.90E-09** | 0.009 | 0.564 | 2 |
| Pancreatic cancer | 1.29 (0.99-1.67) | 5.77E-02 | **1.51 (1.08-2.11)** | **1.64E-02** | 1.93 (0.78-4.77) | 1.60E-01 | -0.01162 | 3.59E-01 | 2.85E-02 | 40 | - | - | - | - | - |
| Gastric ulcer | 1.30 (1.02-1.66) | 3.70E-02 | 1.49 (1.18-1.89) | 9.51E-04 | 1.76 (0.75-4.14) | 2.04E-01 | -0.00864 | 4.72E-01 | 6.42E-12 | 40 | **1.41 (1.13-1.74)** | **3.62E-03** | <0.001 | 0.517 | 3 |
| Rectal cancer | **1.23 (0.89-1.71)** | **2.16E-01** | 1.43 (0.88-2.3) | 1.45E-01 | 1.95 (0.62-6.16) | 2.60E-01 | -0.01310 | 4.16E-01 | 8.78E-01 | 40 | - | - | - | - | - |
| Esophageal cancer | 1.31 (1.01-1.7) | 4.13E-02 | 1.55 (1.18-2.03) | 1.80E-03 | 2.14 (0.87-5.26) | 1.07E-01 | -0.01389 | 2.74E-01 | 1.93E-06 | 40 | **1.36 (1.06-1.75)** | **2.13E-02** | <0.001 | 0.785 | 1 |
| Gastric cancer | 1.28 (1.01-1.62) | 4.12E-02 | 1.39 (1.15-1.69) | 8.22E-04 | 1.60 (0.7-3.67) | 2.73E-01 | -0.00638 | 5.83E-01 | 1.76E-18 | 40 | **1.41 (1.17-1.7)** | **1.15E-03** | <0.001 | 0.719 | 7 |
| Cholecystitis | **1.70 (1.25-2.32)** | **8.04E-04** | 1.83 (1.23-2.74) | 3.02E-03 | 1.64 (0.55-4.91) | 3.80E-01 | 0.00100 | 9.48E-01 | 1.06E-01 | 40 | - | - | - | - | - |
| Cholangiocarcinoma | **1.84 (0.9-3.79)** | **9.68E-02** | 1.89 (0.68-5.24) | 2.23E-01 | 0.44 (0.04-5.42) | 5.25E-01 | 0.04070 | 2.50E-01 | 7.47E-01 | 40 | - | - | - | - | - |
| Diverticular disease | 1.22 (1.02-1.47) | 3.19E-02 | 1.15 (0.95-1.39) | 1.58E-01 | 0.72 (0.39-1.34) | 3.04E-01 | 0.01511 | 8.81E-02 | 8.88E-07 | 40 | **1.27 (1.08-1.48)** | **5.19E-03** | <0.001 | 0.911 | 3 |
| Ulcerative colitis | 0.86 (0.74-1.01) | 6.73E-02 | 0.85 (0.7-1.05) | 1.34E-01 | 0.97 (0.62-1.52) | 8.86E-01 | -0.00180 | 6.00E-01 | 3.21E-28 | 452 | **0.83 (0.72-0.95)** | **9.40E-03** | <0.001 | 0.553 | 8 |
| Crohn's disease | 1.19 (1.02-1.39) | 2.93E-02 | 1.28 (1.06-1.54) | 9.41E-03 | 1.45 (0.92-2.27) | 1.07E-01 | -0.00314 | 3.62E-01 | 1.87E-26 | 452 | **1.13 (0.99-1.29)** | **6.55E-02** | <0.001 | 0.354 | 13 |
| NAFLD | 1.27 (1.12-1.44) | 1.33E-04 | 0.99 (0.84-1.17) | 9.12E-01 | 3.54 (2.06-6.08) | 6.30E-06 | -0.01293 | 1.67E-04 | 9.52E-07 | 474 | **1.50 (1.28-1.74)** | **4.77E-07** | <0.001 | 0.042 | 4 |
| ALD | **1.65 (1.21-2.25)** | **1.51E-03** | 1.83 (1.08-3.11) | 2.44E-02 | 1.32 (0.54-3.25) | 5.42E-01 | 0.00355 | 6.04E-01 | 8.79E-01 | 458 | - | - | - | - | - |
| Primary sclerosing cholangitis | 0.93 (0.69-1.24) | 6.00E-01 | 1.00 (0.65-1.53) | 1.00E+00 | **0.37 (0.14-0.95)** | **4.06E-02** | 0.01412 | 4.74E-02 | 6.75E-02 | 303 | - | - | - | - | - |
| Primary biliary<br>cholangitis | 1.45<br>(1.05-1.99) | 2.29E-02 | 1.70<br>(1.03-2.83) | 3.96E-02 | 1.08<br>(0.43-2.72) | 8.66E-01 | 0.00461 | 5.11E-01 | 1.99E-06 | 310 | **1.54**<br>**(1.16-2.03)** | **2.80E-03** | <0.001 | 0.801 | 4 |

The best causal estimation highlighted in bold. Detailed flow diagram for selecting the best causal estimation is described in Supplementary figure 1.
Odds ratio per 1 standard deviation increase in genetically predicted WC (12.52 cm).
The blank (-) for MR-PRESSO estimation indicates no significant outliers.
SNP, single-nucleotide polymorphism; GERD, gastroesophageal reflux disease; NAFLD, non-alcoholic fatty liver disease; ALD, alcoholic liver disease; OR, odds ratio; CI, confidence interval; $P_h$ , P value for heterogeneity.

### Multivariable MR analysis

After adjustment for the effect of alcohol consumption and smoking in the multivariable MR analysis, the findings were similar to the univariable analysis with a few exceptions. BMI appeared to be significantly associated with acute pancreatitis after adjustment for alcohol consumption with alcohol-adjusted OR 1.27 (95% CI 1.01–1.58). BMI was no longer significantly associated with cholangiocarcinoma and CD (Supplementary Table 5). For WC, the ORs of GERD, pancreatic cancer, gastric cancer, and UC were attenuated to the null after adjustment for alcohol consumption and smoking. WC was no longer associated with gastric ulcer and esophageal cancer after adjustment for alcohol consumption (Supplementary Table 6). Due to the cumulative evidence regarding cholelithiasis as a risk factor for cholecystitis, we assessed the potential mediating effect of cholelithiasis on cholecystitis using multivariable MR. The cholelithiasis-adjusted ORs for cholecystitis were 1.23 (95% CI 0.98– 1.55) and 1.19 (95% CI 0.87–1.63) for BMI and WC, respectively, indicating that the causal effect of adiposity on cholecystitis is likely to be mediated through gallstone formation (Supplementary Table 7). Considering the lack of evidence ensuring the direction of effect among PBC/PSC and UC/CD, we performed a bidirectional MR analysis and identified that UC exerted a causal effect on PSC but not vice versa (Supplementary Table 8). Scatter and funnel plots of 19 GI outcomes are provided in Supplementary Figures 2-5.

## Discussion

This study is the first to explore the causal effect of adiposity on the entire GI system using an MR approach in a large sample size. In total, we incorporated genetic data from over 600,000 European individuals in our analyses. The holistic approach covering the broad GI disorders provides a bird’s-eye view on shared etiological pathways between different GI diseases and offers new insights into organs that are particularly susceptible to increased accumulation of body fat. We identified that increased adiposity was strongly associated with NAFLD, cholecystitis, and cholelithiasis in both BMI and WC measures, which raises concern for the fat vulnerability of hepatobiliary organs. Abdominal obesity measured by WC might be more influential and relevant with a diverse span of GI diseases than simple body size indicated by BMI, supporting a possible pathophysiological role of visceral abdominal fats in the development of GI disorders and cancers.

Given that data from RCTs of interventions against obesity are rare or non-existent, a causal relationship between increased adiposity and health outcomes is extremely difficult to establish.^53^ However, the MR approach enabled the exploration of causal inferences among perplexing epidemiological relationships between obesity and digestive systems using human genetics. From our findings that obesity predominantly elevates the risk of malfunction in the hepatobiliary organs primarily involved in the fat metabolism,^54,55^ we hypothesize that disturbance in lipid metabolism may ground the pathogenetic mechanisms for the obesity-induced hepatobiliary diseases. A possible link between adiposity and liver risks could be the FTO, given that our analysis identified variants associated with the FTO gene being consistently outstanding in scatter plots across diverse hepatobiliary outcomes (Supplementary Tables 9 and 10). It is well established that variants within the FTO region influence body mass and composition phenotypes through functional regulations of IRX3 and IRX5 expression and many other metabolic pathway molecules.^56,57^ Further evidence showed that hepatic FTO is associated with lipid metabolism by reducing fatty acid oxidation and lipolysis and increasing de novo lipogenesis in the liver, and thus the overexpressed hepatic FTO was correlated with increased hepatic fat accumulation.^58-60^ Excessive accumulation of nonesterified fatty acids and triglycerides (TGs) in the liver could lead to cytotoxicity, so-called lipotoxicity, resulting in the development and progression of liver diseases.^61-63^

Interestingly, our results showed that not only NAFLD but also ALD were robustly associated with WC, even after adjustment for alcohol consumption in multivariable MR (OR per 1 SD increase in WC: 1.68, 95% CI: 1.24-2.28, Supplementary Table 6). Given that ALD’s pathogenesis also involves fat deposition in hepatocytes at an early stage,^64^ it is likely that the shared etiology of ALD and NAFLD with respect to adiposity lies in disrupted lipid metabolism that results in abnormal lipid accumulation in the liver. Our result is contextually in concordance with the study by Anstee *et al*. that highlighted shared genetic variants among ALD and NAFLD in the *PNPLA3* gene region for which a substitution in the 148^th^ amino acid (I148M) appeared to promote the hepatic accumulation of TG.^65^

Obesity is associated with increased bile acid (BA) synthesis, which provokes supersaturation of BA in the gallbladder and subsequently gallstone formation (cholelithiasis).^66-68^ Cholecystitis, an inflammation of the gallbladder most often developed by a gallstone,^68,69^ was causally associated with increased adiposity along with cholelithiasis in our results (Figures 2 and 3). The result was in concordance with a previous MR-based phenome-wide association study that reported the causal effect of BMI on cholecystitis.^70^ However, this previous study did not examine the potential mediating effect of cholelithiasis on the development of cholecystitis. To investigate the mediating effect, we conducted multivariable MR analyses and identified that cholecystitis no longer retained a causal relationship with both BMI and WC after adjustment for cholelithiasis (Supplementary Table 7), implying that the causal effect of adiposity on cholecystitis is likely to be mediated through gallstone formation rather than the effect of adiposity per se. Since cholecystitis mediated by cholelithiasis indicated vertical pleiotropy but not horizontal pleiotropy, this mediating effect does not necessarily weaken the assumptions and reliability of our MR results.^13^

Numerous studies have reported that 15-40% of patients with inflammatory bowel disease (IBD) are obese, and an additional 20-40% are overweight.^71,72^ Although grouped together in IBD, CD and UC are known to have heterogeneous etiologies, disease mechanisms, and genetic backgrounds.^73^ Similarly, obesity was shown to be associated with a risk of CD, but not with UC, although results vary across studies.^7,74-76^ To date, findings on the association between adiposity and IBD were derived from observational studies, and thus, the genuine causal relationship remained uncertain. To pinpoint the causal inference, we leveraged genetic instruments as well as a large sample size (>23,000 IBD cases) and confirmed that increases in BMI and WC are suggestively causally linked to an increased risk of CD and reduced risk of UC (Figures 2 and 3). While obesity may be protective against UC, the result should be interpreted with caution given that the estimations were not consistent across diverse MR methods and the relationship between WC and UC changed toward the null findings when adjusted for smoking in multivariable MR (Supplementary Table 6).

PSC and PBC are chronic cholestatic liver diseases characterized by progressive biliary duct damage and fibrosis.^77^ Practical guidance for PBC from the American Association for the Study of Liver Diseases (AASLD)recommended avoiding obesity. However, the advice was given in general terms to patients with PBC, as obesity is generally unfavorable to those with any form of liver disease.^78^ It was challenging to find data on enough people with PSC and PBC for an adequate sample size, and there was a lack of evidence regarding the association between obesity and PSC/PBC. To counter this lack of evidence, we used solid instrumental variables for exposure (Supplementary Tables 11-14) and large consortia data and meta-analyzed GWAS results of PSC/PBC to conduct the first MR analysis.^79,80^ Our findings suggested that obesity is a putative causal risk factor for PBC but not for PSC. Such diverging effects could be explained partially with IBD. Approximately two-thirds of all patients with PSC have comorbid IBD, of whom over 75% had UC.^81,82^ Which occurs first between PSC and UC has long been controversial, and our bidirectional MR results revealed that genetic variants associated with UC significantly increased the risk of PSC but not vice versa (Supplementary Table 8). Given the inverse association of UC with increased adiposity (Figures 2 and 3), we hypothesize that the risk of UC nullifies the effect size of PSC, compromising the risk of PSC while marginally affecting PBC.

A recent consensus statement from the IAS and ICCR Working Group of Visceral Obesity argued that WC was more closely correlated with the absolute quantity of visceral fat compared with other anthropometric indices.^24^ We observed that a larger number of GI phenotypes were robust in association with WC, an indicator of abdominal or central obesity, than with BMI. Besides, the effect sizes of GI outcomes were on average larger with WC than BMI, and the difference was more prominent when comparing the same clinical entity (Figures 2 and 3). Altogether, the pattern may emphasize the more significant influence of fat mass in the visceral cavity over fat distribution in other locations,^24^ which is sensible given the close or even adjacent spatial-anatomical relationship between the abdominal fat pad and organs in the abdomen cavity. One explanation for the greater impact of visceral fat on GI outcomes could be the paracrine action of adipose tissues. Adipose tissues continuously secrete proinflammatory molecules with a marked proportion to the local cell environment.^83^ This could imply that GI organs are influenced by both endocrine and paracrine cytokines, differing from other distant organs that are only affected by endocrine secretions from adipocytes. Moreover, activated paracrine signals via adipokines have been shown to alter the tumor microenvironment in organs closely spaced with the fat pad, partially explaining the pathogenesis of GI cancers.^83^ This mechanism supports our result that GI cancers are more robustly associated with WC than BMI.

### Strength and limitations

This study has several strengths. First, the study adopted a holistic approach covering the entire GI system and identified the putative shared mechanisms and patterns across GI organs influenced by adiposity. Second, the MR design mitigates the bias induced by reverse causality. It is known that the GI tract induces obesity through its contributions to satiety, appetite, absorption of nutrients, and energy balances,^62^ which has long hampered the elucidation of genuine adipose effect on GI outcomes in observational studies. MR analysis uses alleles that are randomly allocated and fixed at conception and thus is unlikely subject to reverse causation.^10^ Third, we used multiple MR models and multivariable analysis to avoid false-positive inferences and further adjust for potential confounders, ensuring careful interpretation of the results.

This study has several limitations. First, the statistical power was not sufficient (<80%) for a few outcomes. Given that insufficient statistical power has been a limitation of MR analysis,^10,11^ we endeavored to maximize the power by using the data with the largest sample size to date and meta-analyzing available cohorts for each GI outcome. Moreover, among robust estimates in our analysis, only gastric cancer was low in power with BMI and WC, implying that the statistical power is less likely to alter our interpretations overall. Second, MR estimates reflect the effect of lifelong exposure.^13,84^ Therefore, our results cannot inform the short-term effect of fat gain on GI outcomes. Lastly, we restricted study samples to individuals of European ancestry to minimize the population stratification bias. This, in turn, prevent our findings from being generalized to other ancestries.

## Conclusion

This comprehensive study identified that adiposity exerted a suggestive causal effect (P<0.05) on the risk of more than half of the GI outcomes and a robust causal effect (Bonferroni threshold P<0.0026 and consistent results across multiple MR methods) on nearly a quarter of the GI outcomes. Hepatobiliary organs (liver, biliary tract, and gallbladder) that are functionally related to fat metabolism were particularly susceptible to increased adiposity. Abdominal obesity measured by WC might be more influential and relevant than BMI with a diverse span of GI disease risks, highlighting a possible pathophysiological role of visceral abdominal fats in the development of GI disorders and cancers.

## Data Availability

All data produced in the present work are contained in the manuscript

